# Comparing the effectiveness of the NHS Health Check and the Polypill Prevention Programmes in the primary prevention of heart attacks and strokes

**DOI:** 10.1101/2023.10.06.23296215

**Authors:** Nicholas J. Wald, Aroon D. Hingorani, Stephen Vale, Jonathan P Bestwick, Joan Morris

## Abstract

**Objective:** To compare the NHS Health Check Programme with the Polypill Prevention Programme in the primary prevention of heart attacks and strokes.

**Design:** Use of published data and methodology to produce flow charts of the two programmes to determine screening performance and heart attacks and strokes prevented.

**Setting:** The UK population.

**Intervention:** The NHS Health Check Programme using a QRisk score on people aged 40-74 to select those eligible for a statin is compared with the Polypill Prevention programme in people aged 50 or more to select people for a combination of a statin and 3 low dose blood pressure lowering agents. In both programmes people had no history of heart attack or stroke.

**Main outcome measures:** In 1000 people the number of heart attacks and strokes prevented in the two programmes.

**Results:** Assuming 100% uptake and adherence to the screening protocol, in every 1000 persons, the NHS Health Check Programme would prevent 287 cases of a heart attack or stroke in individuals who would gain on average about 4 years of life without a heart attack or stroke, the precise gain depending on the extent of treatment for those with raised blood pressure, and 136 would be prescribed statins with no benefit. The corresponding figures for the Polypill Prevention Programme are 316 individuals who would, on average, gain 8 years of life without a heart attack or stroke with 260 prescribed the polypill with no benefit. Based on published estimates of uptake and adherence to of the NHS Health Check programme, only 24 cases per 1000 are currently benefitting instead of 287. This result could be achieved in the Polypill Prevention Programme with just 8% (24/316) of the eligible population taking part.

**Conclusions:** The Polypill Prevention Programme is by design simpler with the potential of preventing many more heart attacks and strokes; even an uptake of 40% would represent a 5-fold greater preventive effect than the NHS Health Check Programme.

## Introduction

Screening in the primary Prevention of future cardiovascular disease is currently based on performing periodic health checks among adults based on clinical history, a limited physical examination including blood pressure measurements and a blood test to measure cholesterol to derive a person’s risk of developing cardiovascular disease. Such screening is conducted in several countries including the UK. The risk assessment was originally based on the results of the US Framingham Study^1^ and later adapted using other data. UK primary care data were used to develop the multi-factor QRisk prediction estimator in the NHS Health Check Programme conducted in England for people who are aged 40 to 74^2^. In the NHS Health Check Programme, QRisk is used to estimate the risk of a cardiovascular disease event over the next 10 years.

Heath Checks, which are the responsibility of local authorities, are conducted every five years by some occurring within and some outside general practices. Persons with a ten-year cardiovascular disease risk of 10% or more, are deemed screen positive. All individuals are offered advice on lifestyle with prescribing of any medication subsequently being left to GPs, with Public Health England advising that a statin is prescribed ‘where lifestyle modification has been ineffecfive or is inappropriate’. If the person’s blood pressure is not thought to be raised, blood pressure lowering medication is not offered.

In 2000, recognising that age as a predictor overwhelms everything else, a simpler method of screening was proposed in a patent application^3^ using age alone without testing or physical examination. This was brought to wider attention in the BMJ in 2003^4–6^ in what has become known as the ‘polypill papers’^7^.

A Polypill Prevention Programme has been offered outside the NHS for over 10 years as a service accessible on www.polypill.com. The Polypill Prevention Programme is directly linked to risk reduction where people aged 50 and older without contraindications, are offered a combined formulation of medicines: a Polypill. This consists of a cholesterol-lowering statin and three low-dose blood pressure lowering medicines to lower both risk factors together *regardless of their starting values*. Randomised trials have demonstrated the value of the polypill versus usual treatment with few side effects^8–12^.

The contrasting approaches of the NHS health check programme and the Polypill Prevention programme raise questions about the future of public health policy on the primary Prevention of cardiovascular disease. First, assuming 100% uptake and adherence to both preventive programmes how do they compare? Second, how does the NHS Health Check Programme perform in practice, where uptake and adherence are unlikely to be 100% and how is this likely to compare with the Polypill Prevention Programme? Third, how should the benefits of the two approaches be quantified and compared? We here answer these questions using evidence published by others and building on our previous work^4,13–16^.

In this study we compare NHS Health Check screening with age alone screening, but the results apply to other multiple risk factor-based screening programmes for the primary Prevention of cardiovascular disease.

## Methods and Results

Published results, were used to construct three flow diagrams to show the screening performance and preventive effect in 1000 persons in the population regardless of age, divided into those that have a future fatal or non-fatal heart attack or stroke over their lifetime (affected) and those that will not have either event (unaffected). One flow diagram is for the UK NHS Health Check Programme, and one is for the Polypill Prevention Programme (Figures 1 and 2 respectively). Both flow diagrams assume complete protocol adherence. The third flow diagram (Figure 3) is for the NHS Health Check programme constructed based on results on uptake and statin use from a published audit of the programme.

**Figure 1:**
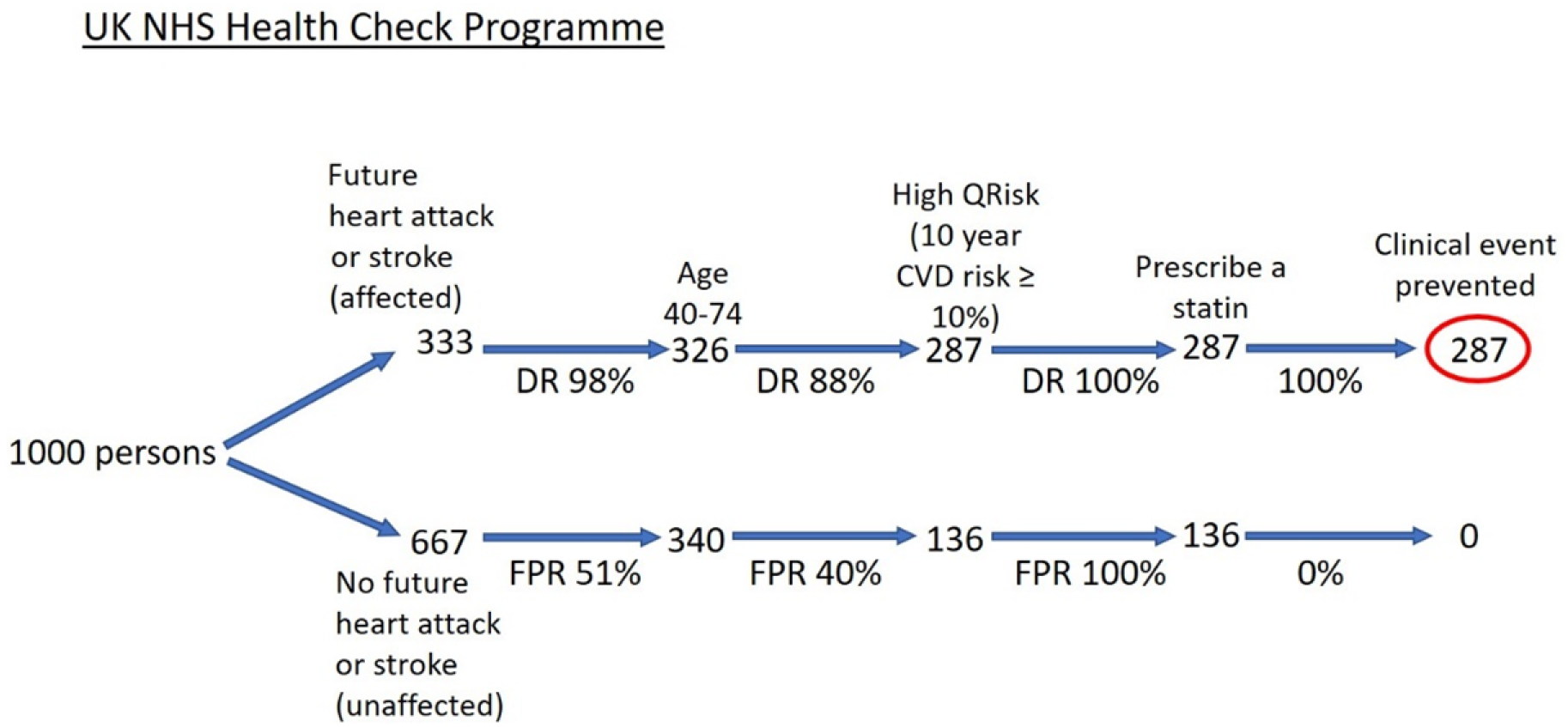
Flow diagram of the NHS Health Check Programme among 1000 people in the population with 100% uptake and adherence.

**Figure 2:**
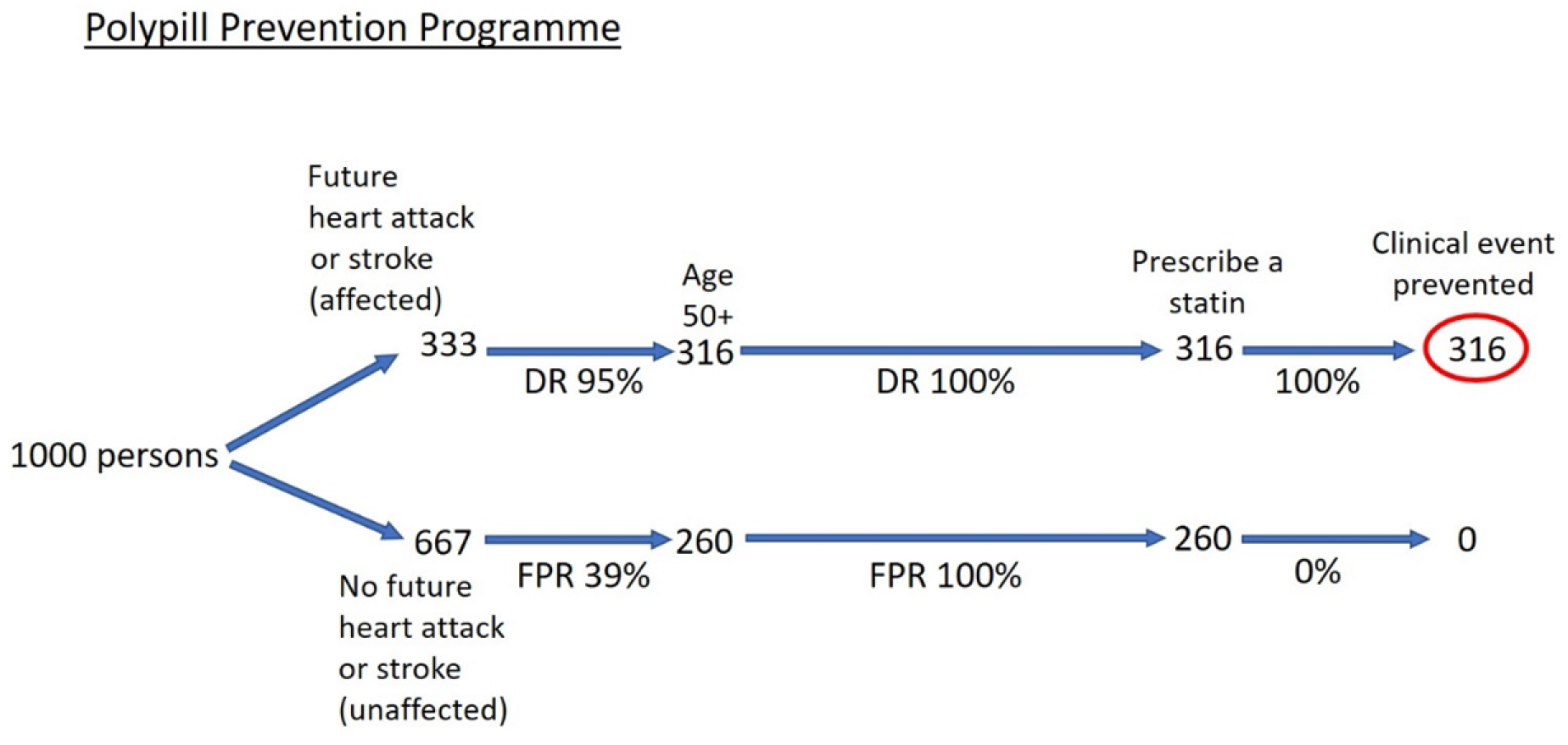
Flow diagram of the Polypill Prevention Programme among 1000 people in the population with 100% uptake and adherence.

**Figure 3:**
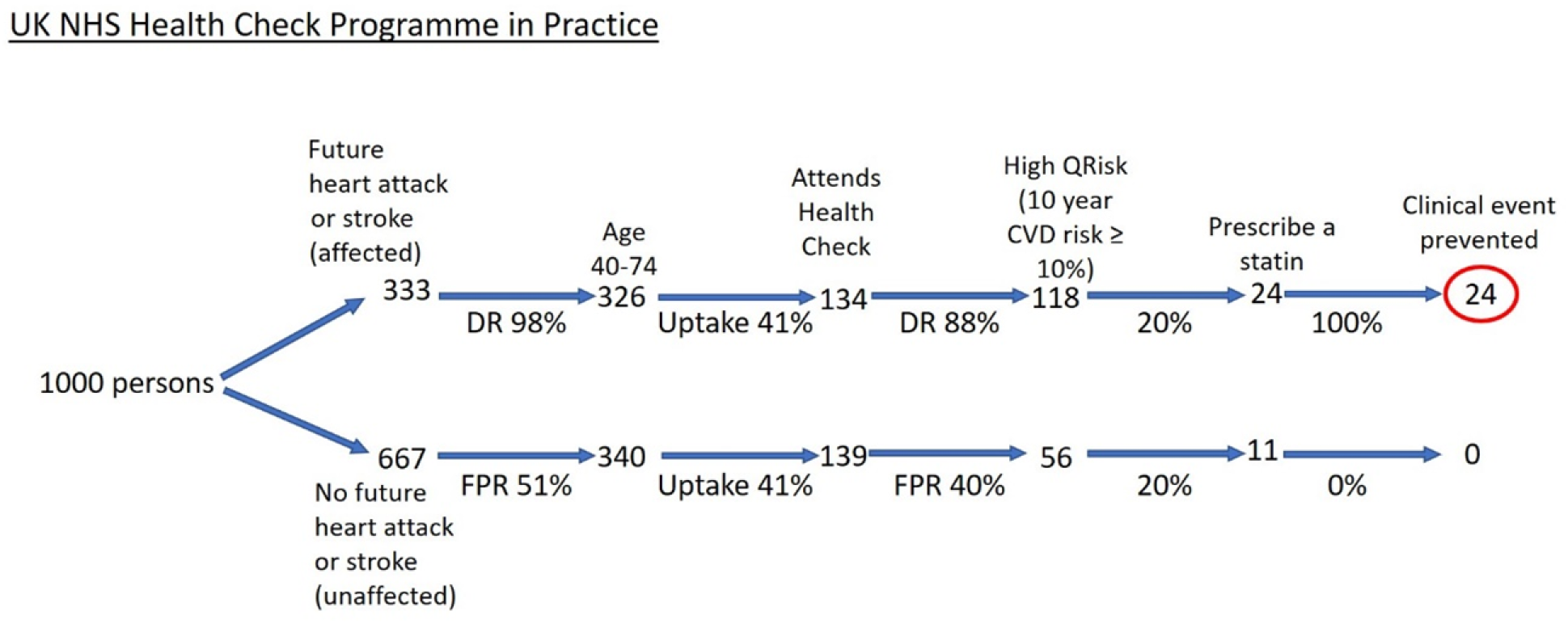
Flow diagram of the NHS Health Check Programme among 1000 people in the population taking account of the observed uptake and statin use.

### NHS Health Check Programme

About 1 in 3 people in the population (333 per 1000) will be affected by a heart attack or stroke over their lifetime and the remainder (667 per 1000) will be unaffected^14^. The first step is identifying people aged 40-74 years as shown in Figure 1. Standard life-table analysis shows that about 98% of affected individuals (326 of 333) and about 51% of unaffected individuals (340 of 667) will be aged 40 or older^17^; the actual estimates will be somewhat less with a 74 year upper limit as specified in the NHS health check programme (eligible people aged 40-74). The second step in the screening process is identifying those with a positive QRisk score, i.e. a ten-year cardiovascular disease risk greater than or equal to 10%. With 5 yearly assessments, using a 10% ten-year cardiovascular disease risk cut-off, the detection rate is 88% (proportion of individuals with a first cardiovascular disease event in the next 10 years with positive screening results) and the false positive rate is 40% (proportion of individuals who do not have a first CVD event in the next 10 years with positive screening results). This yields 287 true positives (88% of 326) and 136 false positives (40% of 340). These estimates are based on the Framingham risk score^13^ which has similar performance to QRisk. The third, and final step, is prescribing a statin to QRisk positive individuals.

Figure 1 shows that in the NHS Health Check Programme all 287 true positives receive a statin and will, as a result, have a heart attack or stroke entirely prevented or delayed, since medication with a statin will have a preventive effect in all who take it, albeit with a variable preventive period adopting the holisfic model described by Wald and Morris in 2014^14^. The holistic model shows that on average, a gain of 4 years of life without a heart attack or stroke could be achieved with a statin, which may be somewhat longer depending on the use of blood pressure lowering medications being prescribed in the NHS Health Check Programme if the blood pressure were considered to be high.

### Polypill Prevention Programme

Figure 2 shows, in the same way as Figure 1, the results of the Polypill Prevention Programme which uses age alone in selecting people for preventive medication. Among persons in the affected group shown in the Figure, standard life-table analysis^14,17^ indicates that 95% of first heart attacks and stroke will be found in those aged 50 and over (not much less than the 98% found in those aged 40 and over), yielding 316 true positives. In the unaffected group, life-table analysis indicates that 39% of all people who do not have a first heart attack or stroke will be found in those aged 50 and over yielding 260 false positives. All 576 individuals (true and false positives) are prescribed a polypill including a statin and three low-dose blood pressure medicines with 316 clinical events prevented or delayed gaining, on average, 8 years of life without a heart attack or stroke. In contrast with the NHS Health Check Programme, the Polypill Prevention Programme includes blood pressure lowering medication for everyone.

### Implementation of the NHS Health Check Programme

NHS Health Checks, started in England in 2009, have been audited in England and the results are likely to apply for the UK as a whole. Figure 3 shows a flow diagram taking account of an audited 41% uptake^18^ and a 20% use of statins in QRisk positive individuals^16^. A 2021 audit yielded lower uptake and adherence estimates^15^. Using the estimates of 41% and 20% respectively, out of 1000 people, 24 cases would be prevented, (Figure 3) compared to 287 with full uptake and everyone with a positive QRisk receiving a statin (Figure 1), i.e., 92% fewer.

### Comparison of the Programmes

The Table shows the differences in the two screening programmes with 100% uptake and protocol adherence. In every 1000 people the Polypill Programme prevents 29 more heart attacks and strokes (316 minus 287) than the NHS Health Check Programme with 124 more false positives (260 minus 136) and *no clinic visits or blood tests*. But in practice, uptake and protocol adherence of the Health Check Programme is far from 100%. Despite the uptake in a Polypill Prevention Programme not being known, using the audit results of the NHS Health Check Programme indicates that uptake in a Polypill Programme would have to be only 8% (24/316, from Figures 2 and 3) to achieve a better outcome in terms of clinical events prevented.

**Table 1.** Comparison of NHS Health Check Programme and Polypill Programme among 1000 people in the population.

|  | NHS Health Check Programme | Polypill Prevention Programme |
| --- | --- | --- |
| 1. (i) In every 1000 people: number of heart attacks and strokes prevented | 287 | 316 |
| (ii) Receiving medication who would or would not have had a heart attack or stroke | 136 | 260 |
| (iii) Number of clinic visits | 7000* | 0 |
| 2. No. of screening steps | 3<br>(age, QRisk, prescribe a statin) | 1<br>(age only) |
| 3. Preventive medication | Statin with or without blood pressure lowering medication | Statin and low-dose blood pressure medication (polypill) |
| 4. Average years of life gained without a heart attack or stroke | 4** | 8 |
| 5. Uptake and adherence | Very low | Unknown |

\*1000 x 7 for 7 five yearly 'Health Checks'
\*\* with statin only

## Discussion

The flow charts in Figures 1 and 2 show that with complete uptake and adherence, the Polypill Prevention Programme prevents more heart attacks and strokes than the NHS Health Check Programme with no clinic visits or blood tests. Figure 3 shows that the NHS Health Check programme has a very limited effect and consideration needs to be given to replace it with a simpler more effective polypill approach. In making such a switch in policy several issues, covered below, need to be considered.

### NHS Health Check Approach

The NHS Health Check Programme is a three-step screening programme. First people are selected based on age (40-74). Then tests are performed to determine a person’s ten-year risk of a cardiovascular disease event. This decreases the false positive rate, but also decreases the detection rate. Third, a clinical decision is made on whether a statin is prescribed, possibly only after attempting to reduce risk through diet and lifestyle, and separately, based on blood pressure measurements, a clinical decision may be made on whether a blood pressure lowering medication is prescribed. However, where a preventive intervention is safe and affordable, as is the case with statins and blood pressure lowering medications^5,6,19^, the balance should be in favour of a simple approach that increases detection and disease Prevention employing a policy that directly offers access to preventive medication.

Aside from the low uptake, and adherence to the NHS Health Check Programme there is another weakness. The aim of preventive medication should be to reduce the risk of cardiovascular disease as much as possible which is achieved by lowering blood pressure *and* LDL cholesterol together, not just one or the other if either are thought to be abnormally high, and to do so *regardless of the starting level* because there is no practical blood pressure threshold below which there is no further reduction in risk^20–22^.

A ten-year risk of a heart attack or stroke equal to or greater than a given percentage (say ≥ 10%) is too limited for assessing risk or benefit in chronic disease Prevention, where the risk of disease is lifelong and requires lifelong preventive intervention. It also does not adequately define the benefit from preventive intervention. Lifetime risk and lifetime benefit is what is relevant. A further consideration relates to missed preventable cases in the NHS Health Check Programme. If a person has a positive QRisk assessment (i.e., ≥10% ten-year risk) preventive medication is offered. Once started it will, presumably, be taken for the rest of that person’s life. From about age 74 almost everyone will be QRisk screen positive. The effect of the NHS Health Check Programme will be to miss the opportunity to prevent some heart attacks and strokes at younger ages that could have been prevented with an age alone policy. For example, a 60-year-old could have a negative assessment, but on an age-alone policy would be positive and eligible for preventive medication.

There is minimal benefit in starting preventive medication below age 50 as is the case in the NHS Health Check Programme. Selecting age 50 as the age a polypill is offered is a policy judgement that may vary from country to country depending on the age distribution of heart attacks and strokes, cost and affordability. This is because there are very few people below age 50 who have a positive QRisk (reflecting the fact that age is the most important determinant of risk), very few events occur between age 40 and 50, and the full effect on risk reduction from blood pressure and LDL cholesterol reduction is achieved after only about 3 years^5^.

In 2014 the NHS Health Check Programme lowered the 10-year cardiovascular risk threshold from 20% to 10%, but this led to little change in practice; the average calculated risk of all people who were started on statins was about 21% before the guideline was changed and about 20% after^16^. This calls into question what the programme achieves and indicates that the policy was not followed.

In a survey of 1.4 million people registered with 248 UK general practices, 73% of individuals initiated on a statin did not have a QRisk score recorded at any time (37,215/50,940)^23^. On this basis alone it can be concluded that a multi-factor screening ‘tool’, such as QRisk, plays only a minor role in influencing the decision to start people on a statin.

Importantly the results in Figure 3 show that the Health Check Programme has a very small effect, with the unavoidable conclusion that it should be replaced by a more effecfive programme.

### Polypill Prevention Approach

Apart from already having had a cardiovascular event, the overriding influence on a person’s future risk of a cardiovascular event is that person’s age. From youth to old age the risk of cardiovascular events doubles every seven to eight years^24^. Other risk factors, like blood pressure and LDL cholesterol, although causal and reversible, are poor predictors of disease^25^. It follows that in the primary Prevention of cardiovascular disease, age alone can be used in a once only screening enquiry. There is only a marginal improvement in screening performance over age alone from adding causal risk factors^13^. A person’s age is already available from that person’s medical records. Age alone is the screening test that determines eligibility for preventive medication that can then be offered in the absence of specific medical contraindications. Indeed, while selection on age is a critical first screening test for almost all screening programmes it is not recognised as such, but its importance is shown in the Figures as a critical step even in the NHS Health Check screening pathway.

Appropriately formulated with a statin and low dose blood pressure medications a polypill has a low incidence of side effects. In a crossover trial there were no withdrawals due to side effects^19^. Comprehensive analysis on statins indicated side effects were rare^26^. The use of blood pressure medication at low doses in randomised trials showed a very low rate of side effects^6^ and a reduction of one third in headache^27^ which was confirmed in polypill participants (https://www.polypill.com/Home/Headaches). Importantly, side effects are reversible on stopping treatment and vastly outweighed by the benefit. In the UK Polypill Prevention Programme less than 1% of the participants discontinued on account of side effects, all of which were minor, and not necessarily related to taking the polypill.

High cost and potential harm are reasons to limit the offer of a medical intervention to screen-positive individuals selected based on precisely determined high risk estimation. But this is not the case if Prevention is inexpensive and safe as it is with a polypill, justifying the conclusion that screening can be based on age alone, with a starting age just before heart attacks and stroke become a significant disease burden in the population. In such a situation Prevention is better than prediction.

The polypill approach is a public health primary cardiovascular disease Prevention programme that is based on age alone and the use of an optimally formulated polypill. It has advantages over a multiple risk factor screening approach, such as the NHS Health Check Programme that focuses on disease prediction and leaves preventive treatment to doctors. The Polypill approach is simpler, saves practitioner time and avoids routinely carrying out periodic blood pressure measurements and blood tests. Medical practices could identify people once they have reached age 50 and offer them, by email/post or text message, a polypill. The polypill would be a standard formulation very much like a vaccine and could be dispensed by local pharmacies. The ethos would be to take the medication to avoid becoming a patient, not because one had become one, recognising that if a first heart attack or stroke is prevented there is no second one to prevent. Costs would be largely limited to producing the polypill and making it accessible to the public, not adopting an expensive complex protocol with repeated measurements every 5 years to estimate the risk of disease: The NHS Health Check has been shown to take typically 20-30 minutes for each assessment^28^ and a report in 2015 estimated that the annual cost was £450million^29^.

Determining who is offered a polypill can be assessed by asking a few simple on-line questions as is done in the Polypill Prevention Programme (www.polypill.com). Such a service could be scaled up and offered by the NHS and other health care providers throughout the world. The service could be audited to determine what proportion of people offered the polypill take it up. There would be little financial waste because the cost would be approximately directly proportional to the uptake. Sample surveys of individuals could be conducted to determine the LDL cholesterol and blood pressure levels in participants in the Prevention programme and in non-participants. A well-managed Polypill Prevention Programme conducted at scale would be an effective, low cost, safe medical intervention that would, to advantage, replace the NHS Health Check Programme. As well as the health benefits^14^ the programme would be cost effective^30^. Uptake and adherence are important. Figures 1 and 2 assume a 100% uptake and medication adherence to the two programmes but in practice this will be less. Figure 3 shows the practical effect of the NHS Health Check Programme. Comparable estimates for uptake and adherence are not available for a Polypill Prevention Programme because the programme in the UK is based on individuals being aware of the service and choosing to become a participant and therefore, not representative of what would happen in the general population. However, the effectiveness of a Polypill Programme is likely to be significantly greater than the NHS Health Check Programme for three reasons. Firstly, the uptake and adherence in a Polypill Programme would only have to be as low as 8% to be better than the NHS Health Check Programme. Secondly, a polypill programme requires little or no inconvenience to people offered such a service through their family doctor or community pharmacy on reaching the age of 50. Thirdly, it has been shown, in several studies that are summarised in a meta-analysis of randomised trials that polypills, improve adherence to treatment over usual treatment^31^.

In May 2023, NICE issued updated guidance that recommends the use of statins in those who have a ten-year QRISK3 score of 10% or more, but not to rule out statin treatment ‘because the person’s ten-year QRISK3 score is less than 10% if they have an informed preference for taking a statin or there is a concern that risk may be underestimated’^32^. The guidance, however, is silent on concurrent blood pressure reduction which has as large an effect on stroke Prevention as statins have on preventing heart attacks. Notwithstanding this omission, the risk thresholds for preventive medication have fallen over time, from 20% to 10% 10-year risk – effectively moving towards a simple age-alone polypill based programme. In this context it is significant that in April 2023 the Hewitt Review^33^, commissioned by the UK Government, endorsed the view that the primary Prevention of cardiovascular disease was a national public health priority. A milestone in the acceptance of the polypill was made in July 2023 when the World Health Organization listed a polypill for the primary and secondary Prevention of cardiovascular disease on its list of essential medicines^34^.

### Quantifying the health benefits of cardiovascular disease Prevention

A ten-year risk of a heart attack or stroke equal to or greater than a given percentage (say ≥ 10%) is not an appropriate metric to summarise the health benefits of chronic diseases Prevention. The ten-year risk period is arbitrary, while the risk of disease is lifelong, requiring lifelong preventive medication and the benefit from preventive intervention is inadequately defined. The limitations arise because the metric adopts the ‘reductionist’ model of assessing benefit, in which people either do or do not benefit^14^. It is a categorical measure that ignores the preventive effect of delaying the occurrence of a heart attack or stroke. In the reductionist model the relative risk reduction is used to identify two separate groups, one group consisting of the number of people who have a clinical event that the intervention is designed to prevent and another group that does not; one group experiences all benefit and the other experiences no benefit at all. The reductionist model, however, is not appropriate for the Prevention of a chronic disease, such as ischaemic heart disease in which clinical events arise from the disease over time. In expectation, the clinical events are delayed to a variable extent in everyone who would have an ischaemic heart disease event when not taking preventive treatment.

An alternative approach that overcomes the limitations of the reductionist model is the holistic model^14^ which is used in the Figures. In the holistic model everyone who would have had a heart attack or stroke in the absence of treatment benefits because blood pressure and/or LDL cholesterol lowering treatment will always delay such an event, albeit to a varying extent; it recognises that a clinical event delayed is a preventive benefit. The reductionist model counts only people who do not have a heart attack or stroke as benefitting, often without taking account of time gained. Two metrics summarise the quantitative benefit using the holistic model. The first is the lifetime probability of a person benefitting from the intervention, which, with preventive medication starting at age 50, will benefit 1 in 3 people, which is approximately the proportion of people in the population that will have a heart attack or stroke because heart attacks and strokes are rare under age 50. The second is, among those who benefit, the average gain in life without a heart attack or stroke, which, for an appropriately formulated polypill, is 8 years. We are aware of no other public health measure that would currently have as large an impact on the primary Prevention of disease as the Polypill Prevention Programme. Using a statin alone, about 4 years of life gained without a heart attack or stroke. It is necessary to assess lifetime risk with and without the preventive medication and express the benefit in terms of lifetime gained rather than the number of cases that do or do not have a heart attack or stroke over, say, a ten-year period. Lifetime gained is an essential measure arrived from the holistic model but is not essential in the reductionist model.

## Conclusion

The NHS Health Check Programme is less effective than the Polypill Prevention Programme in the primary Prevention of heart attacks and strokes. Replacing the NHS Health Check Programme, and similar programmes around the world, with Polypill Prevention Programmes would secure significant health benefits to individuals and populations.

### Post-script on the Polypill Approach

There is widespread recognition that more needs to be done in the primary Prevention of heart attacks and strokes which remain a leading cause of premature death and disability throughout the world. The polypill approach to primary Prevention would have a major impact. Despite the polypill approach being set out and quantified in detail over 20 years ago, positive clinical trial results, and the evidence submitted to the World Health Organisation which resulted in the polypill being added to its list of essential medicines in 2023^35^ (all cited in our paper), no large scale polypill primary cardiovascular disease Prevention programme has been implemented. This post-script to our paper discusses the reluctance to consider and adopt the polypill in the primary Prevention of heart attacks and strokes, the formulation of the polypill in current use, its prescription as an unlicensed medicine and what can be done to facilitate its use as a routine public health service.

### Reluctance to adopt the polypill approach

There has been concern that the polypill approach has not been adopted in spite of its benefits ^7,36,37^. There are a number of reasons that may account for this.

First, the magnitude of the benefit may not have been widely recognized. The flow chart analysis in the accompanying paper sets out and clarifies the practical outcome benefits of the polypill approach compared to the NHS Health Check Programme in England in a way that has not previously been done. Importantly it reveals the shortcomings of the NHS Health Check programme in preventing heart attacks and strokes.

Second, the reluctance may reflect a conservative attitude to the use of medicines for the Prevention of disease while accepting the same medicine for the treatment of patients with disease. Concerns over “medicalisation” are misplaced; the Polypill Prevention Programme has the opposite effect, in that it prevents a person becoming a patient in the same way that people take anti-malarial medication to prevent contracting malaria. The issue is discussed in former *British Medical Journal* Editor Richard Smith’s blog^38^. In contrast the NHS Health Check Programme makes a patient out of everyone selected for a statin.

Third, there is an unjustified perception that more scientific research is needed on the polypill approach. There is already sufficient evidence that a well formulated polypill is safe and greatly reduces blood pressure and LDL cholesterol which in turn prevents heart attacks and strokes^19^. However unjustified the percepfion, there is a need to engage with, and persuade policy makers of the advantages of the polypill approach.

Fourth, clinical guidelines have become increasingly focused on multifactorial risk prediction models and the addition of new predictors. This focus on prediction rather than Prevention has added complexity with recent updates to existing guidelines being incremental instead of taking a fresh look at the whole approach to the Prevention of cardiovascular disease.

Fifth, and relatedly, professionals tend to overlook the fact that age is the most powerful predictor of cardiovascular disease, with little discrimination added by the addition of information from causal risk factors such as blood pressure and LDL cholesterol, or from ‘novel’ biomarkers.

Sixth, there is little financial reward to the pharmaceutical industry in conducting expensive trials to obtain a primary Prevention licence for a polypill which consists of generic medicines some at low non-standard doses. The use of a polypill as a ‘special’ medication is not attractive to the pharmaceutical industry because, although ‘specials’ are regulated, they are not licensed and therefore cannot be marketed.

Seventh, regulations to obtain a product license lack flexibility, tend to rely exclusively on randomised trials performed in the population of intended use, (a major challenge in primary Prevention) and are not proportionate to the evidence on benefit and hazards that is already available from trials and cohort studies that are not performed in the population of intended use.

Each of these issues can and should be addressed so that effective and safe medicines for disease Prevention like the polypill are readily available at low cost.

### Formulation of the polypill in current use

An appropriate and validated preventive medication for cardiovascular disease comprises a polypill that combines a statin and three low-dose blood pressure reducing agents from different classes, preferably formulated in a single capsule or tablet^4^. Such a multiple-low-dose approach minimises any side-effects of the blood pressure lowering medicines. As a matter of routine practice, if a statin is indicated, so should a blood pressure lowering medication and *vice versa* because the aim is disease Prevention, not to ‘normalise’ blood pressure or LDL cholesterol when ‘normal’ levels in the population are, on average, too high. The benefit of the NHS Health Check programme is constrained by not routinely prescribing blood pressure lowering medications in people selected for prescription of a statin. Only about an average of 4 years of life are gained without a heart attack or stroke using a statin alone among the one in three people who, in the absence of preventive action, would be affected by heart attack or stroke over their lifetime, instead of about 8 years if both a statin and blood pressure lowering medications are prescribed, calculated using standard life-table methods^14,17^. A pill that contains both a statin and three low dose blood pressure medications ensures that all are used together^39^. The formulation of the polypill used in the UK Polypill Prevention Programme (polypill.com) is: rosuvastatin 10mg, hydrochlorothiazide 12.5mg, amlodipine 2.5mg, losartan 25mg.

Aspirin could be considered as an additional component in a polypill, with a modest added benefit in preventing cardiovascular disease *and* a reduction the incidence of colon cancer and possible other cancer^40–42^. Although aspirin increases the risk of a gastric bleed this is rarely fatal and the balance is in favour of aspirin being used with a polypill. It is less certain whether folic acid, a suggested option in the original polypill proposal, should also be taken. Randomized trial evidence suggests that folic acid has a benefit in the Prevention of stroke^43^ but has not shown a benefit in the secondary Prevention of heart attack^44^. This may be because the routine use of low-dose aspirin in the secondary Prevention of cardiovascular disease has an anti-platelet effect that is not enhanced by the addition of folic acid^45^. If aspirin is not used there is probably an additional benefit in taking folic acid; 0.8mg per day is an appropriate regime daily dose because it has a maximal effect in lowering serum homocysteine^46^.

### The Polypill as an unlicensed medicine

The polypill prescribed in the Polypill Prevention Programme is an unlicensed medicine but this is not a barrier, indeed it has several advantages. The UK regulations covering unlicensed medicines are particularly suitable for formulations that come under the general term “polypill” and this is likely to be the same in other countries. An unlicensed medicine cannot be specifically promoted; it can however be formulated and prescribed to fit a special need that is not covered by a licensed formulation. The cost of producing and conducting a randomized trial of a polypill with particular components at specific doses to show that each component exerts an independent effect in preventing or treating a particular disorder, or set of disorders, is prohibitive. There is no commercial justification for such a trial when individual licensed preparations can be prescribed separately with some tablets split to achieve the desired dose. The difficulty is magnified enormously when the aim is the primary Prevention of heart attacks and strokes because of the much greater number of people needed in a trial to achieve statistical power. There is also the advantage that the composition and dose of a formulation can be varied; if it were licensed it would require fresh trials. Given that there is no need to advertise specific polypill formulations, there is no need to license them. There is no reason why the current service on polypill.com should not be done on a large scale so it is available on a population wide basis with the cost savings arising from the economies of scale. The large scale use of an unlicensed polypill is entirely possible but may face challenges. Regulators would need to see this as a positive public health opportunity and public education would be needed to explain that ‘unlicensed’ does not mean unregulated or substandard. Meeting these challenges should not be a reason for the NHS and other health services to further delay the implementation of the polypill approach as a national preventive service.

### Next steps

The reluctance to adopt the polypill approach in over two decades since it was first described indicates that more needs to be done before policy makers are willing to implement change. To this end it would be appropriate to conduct a large scale project of implementing a Polypill Prevention Programme that was compared with current practice. A simple low-cost cluster randomisation design could be adopted, with integrated health systems being allocated to the polypill approach or usual practice, without the need for individual consent. Clinical outcomes could be sought using record linkage to electronic medical data. Health records could also be used retrieve blood pressure and LDL cholesterol measurements and determine the differences between the polypill and current practice. This implementation project would provide direct evidence that could not be reasonably ignored.

## Disclosure

NW is a Director of Polypill Ltd which runs the UK Polypill Prevention Programme. ADH is a member of the Advisory Group for the Industrial Strategy Challenge Fund Accelerating Detection of Disease Challenge, and a co-opted member of the National Institute for Health and Care Excellence Guideline update group for Cardiovascular disease: risk assessment and reduction, including lipid modification, CG181.

## Data Availability

All data produced in the present work are contained in the manuscript

## Acknowledgements

We thank Richard Smith and Jeff Aronson for their helpful comments on drafts of the paper.

## Appendix

Source of estimates used in the Figures:

Figure 1

The estimate that 333 people of the initial 1000 will be affected with a heart attack or stroke over their lifetime and 667 will be unaffected comes from Reference 14. The estimate that approximately 98% of affected individuals (326) and 51% of unaffected individuals (340) will be aged 40 or more years and so eligible for the NHS Health Check comes from Reference 17. Of these, approximately 88% of affected individuals (287) and 40% of unaffected individuals (136) aged 40 to 74 will have a cardiovascular disease risk greater than 10% (estimated from Reference 13). All these people will be prescribed a statin and so 287 will have a clinical event prevented.

Figure 2

As Figure 1 but with approximately 95% of affected individuals (316) and 39% of unaffected individuals (260) aged 50 or more (from Reference 17) and so eligible to join the Programme. These people will be prescribed a Polypill consisting of a statin and three low-dose blood pressure lowering medicines thus preventing 316 clinical events.

Figure 3

As Figure 3 but with a 41% uptake (NHS digital) of the NHS Health Check and 20% of QRisk positive individuals receiving a statin (from Reference 16), thus preventing 24 clinical events.

## Notes

### Funding Statement

This study did not receive any funding.

